# Computer vision, causal inference and public health modelling approaches to generate evidence on the impacts of urban planning in non-communicable disease and health inequalities in UK and Australian cities: A proposed collaborative approach

**DOI:** 10.1101/2023.04.18.23288757

**Authors:** Ruth F. Hunter, Leandro Garcia, Mark Stevenson, Kerry Nice, Jasper S. Wijnands, Frank Kee, Geraint Ellis, Neil Anderson, Sachith Seneviratne, Mehdi Moeinaddini, Branislava Godic, Selin Akaraci, Jason Thompson

## Abstract

**Background:** Given that the majority of the world’s population live in cities, it is essential to global health efforts that we design them in ways that both reduce non-communicable diseases (NCDs) risk and that facilitate adoption and maintenance of healthy lifestyles. Current approaches tend to focus on the relationship between urban design-related factors that affect health at the local or neighbourhood level but few studies have explored this relationship both within and across entire cities, nor explored the causal pathways between urban-designed related factors and NCDs. The aim of this research program is to use computer vision, causal inference, and public health modelling methods for understanding the causal relationship between urban design and health at the neighbourhood level, and to explore intervention approaches at the city scale.

**Methods:** Phase 1 will use machine learning and computer vision techniques to analyse gridded, local-level aerial images (with an optical resolution of <20cm), of all UK and Australian cities with populations over 100,000. It will identify a variety of urban features within these images and derive associations between them and NCD incidence and risk factors identified through location-based health surveys. Phase 2, using data from prospective health cohorts and linked objective built environment data, will apply Bayesian networks to investigate the possible causal pathways between built environment, lifestyle factors, and NCD incidence. Phase 3 will estimate the health impacts of actionable changes in urban design. Using health impact assessment modelling, we will calculate the NCD burden that could be prevented if cities were to adopt urban features of healthier counterparts. A similar approach will be applied on finer-grained scale within all case study cities, enabling assessment of health impacts of changes in individual locations. Phase 4 will develop an interactive web-based toolkit to enable urban designers, planners and policymakers to inform the decision-making cycle, co-designed with intended users involving participatory workshops.

**Discussion:** We use state-of-the-art approaches to: (i) generate evidence on the impacts of urban planning and design in NCDs and health inequalities in UK and Australian cities, and (ii) provide stakeholders with tools for advocacy and designing healthier cities.

**Trial registration:** Not applicable.

## INTRODUCTION

The world has seen a rapid population increase of 1 billion since 2005 and 2 billion since 1993 to a current population of 7.3 billion. Most of this growth has been in cities [1]. Managing city population growth across high-, middle- and low-income countries is a significant global public health challenge. Safe and healthy food, clean water, clean air, safe housing, transport, and healthy social relationships are among the most basic of human needs. But even in locations of comparatively high economic development, such as the United Kingdom (UK) and Australia, large areas exist where urban design contributes to increased incidence of known non-communicable diseases (NCDs) associated with risk factors such as physical inactivity, air pollution, poor access to public transport, increased access to tobacco and alcohol, inequitable access to nutrient-poor foods, and social disconnection, leading to adverse health and disease outcomes [e.g. 2-9].

Many NCDs are associated with factors related to lifestyles that city design and modern urban living generates [10]. City design can contribute to the risk of type 2 diabetes mellitus, cardiovascular disease, chronic respiratory disease, some cancers, mental illness (commonly existing as multi-morbidities due to shared risk factors and interacting biological mechanisms [11]), in addition to road injury. Despite their relative wealth, Australia and the UK are not immune, having experienced among the greatest global increases in rates of overweight and obesity leading to the development of NCDs and metabolic diseases since the 1980s [12,13]. The combined effect of air pollution, poor diet, high sugar intake, physical inactivity, smoking, and other modifiable lifestyle factors, driven in part by the reinforcing influence of low-density, car and fossil-fuel dependent cities, pose serious barriers to change. This is in addition to the toll from serious injuries and deaths, especially among active transport users namely, pedestrians and cyclists who are otherwise acting healthily [14,15]. This highlights the need for urban design to consider the totality of population health gains when designing new areas or interventions [16]. For example, urban design-related factors such as density, mixed land use, connectivity and destination accessibility affect behaviours such as physical activity and travel behaviour including driving, public transport usage, walking and cycling [17-25]. These behaviours can contribute to health outcomes directly [26-29] and indirectly, from air pollution, noise levels and urban heat islands [30-31]. NCDs have complex causal mechanisms characterised by multiple aetiologies and factors, and are influenced by an individual’s and their peers’ behaviour along with the external urban and socio-economic environment [32-33]. Therefore, a deeper understanding of the features of urban design that promote healthy behaviour, and the possible causal pathways, can lead us toward a better understanding of principles that could be incorporated into city planning for better population health.

Furthermore, NCDs are socially patterned. A number of Sustainable Development Goals (SDGs) set targets that relate to the reduction of health inequalities including health and wellbeing for all (SDG 3) and a reduction of inequalities within and between countries (SDG 10). The interaction between inequalities and NCDs are complex: low socioeconomic status increases the risk of NCDs, chronic ill-health and associated costs. The consequent limitations in capacity to work can, in turn, reduce household income. Compelling evidence from 283 studies supports a positive association between low-income, low socioeconomic status, or low educational status and NCDs [34]. Good public health is a key driver in the SDGs, and reduction of health inequalities and NCDs should become key in the promotion of the overall SDG agenda. A sustained reduction of general inequalities in income status within and between countries would enhance worldwide equality in health. Niessen and colleagues [34] argue that to end poverty through elimination of its causes, actions to tackle NCDs should be included in the development agenda, with a particular focus on mitigating social, health and climate shocks that worsen the already vulnerable socioeconomic condition and health status of the poor. This may be partly achievable through better and more equitable urban design (SDG 11) [35], using novel, scalable methods that can handle the complexities of urban life (i.e., complex interactions between people and places in our towns and cities).

New methods in computer vision, artificial intelligence (AI) and complexity science (including computational social science) now enable researchers to go beyond what has previously been possible in understanding the relationship between urban design and NCDs at scale [36]. Researchers and policymakers can avoid inefficient methods of data collection that use and rely upon individual city administrative sources, and instead harness consistently collected, massive urban datasets on a global-scale using remote sensing such as satellite and aerial imagery. These datasets contain millions of images related to urban form from cities around the world. Streamlined processes and pipelines that ingest data in the form of images, and harness advances in machine learning and computer vision can now be utilised to discern (i) individual features that exist within urban areas, (ii) neighbourhood types featuring combinations of urban features (iii) global city typologies that group cities according to similar urban characteristics [36], and (iv) individual city fingerprints, that create comparable fine-scaled, block-level representations of urban morphologies across individual cities [37]. Further, these can be linked with processes for producing AI-driven urban design solutions using generative adversarial networks, which create images that combine the structure of existing low performance (e.g., poorly designed and unhealthy) urban areas with the design of high performance areas, enabling new urban futures to be imagined [38,39]. These methods enable a virtually limitless representation of cities to be included in analyses, supporting a fine-grained analysis of morphological differences at the block or neighbourhood level to determine how urban form is associated with population health risks and outcomes [40].

While standard research methods using geographic information systems (GISs) and largely linear statistical methods have brought great advances in our understanding of the intersection between urban design and health, their origins in experimental health research sees them better suited to smaller scale interventions, samples, or problems of defined and simpler natures [41] than those that arise from more complex interactions between cities, people, and health. This research program will therefore exploit more contemporary, advanced methods as described above to overcome identified issues that have led contemporary city scientists and public health researchers to call for change [10,42-45]. The proposed research extends our current approaches by integrating longitudinal data with unique spatially derived data enabling, for the first time to explore the complex interactions of NCDs at a city-scale.

Objectives of the research program include:

1. Utilising new methods in computer vision and artificial intelligence to explore the association between urban design, city types, and NCDs in UK and Australian cities.
2. Investigate how inter- and intra-city urban design disparities are associated with inequalities in incidence and prevalence of NCDs.
3. Combine large cohort and GIS data to prospectively investigate the possible causal pathways between urban design, NCD risk factors, and NCD incidence.
4. Estimate improvements in the burden of NCDs that could be achieved through actionable changes to the built environment at different scales.
5. Design and develop an accessible, web-based toolkit tailored for use by urban designers, planners, policymakers and the broader public.

## METHODS AND ANALYSIS

The research program has four phases that aim to: (i) investigate the relationship between urban design and NCDs at the: a) neighbourhood level, and b) city level using a combination of methods that bring together big data, global image sources, and population-based health data; (ii) explore the possible causal pathways between urban design related factors, NCD risk factors and NCD incidence; (iii) estimate the potential reduction in NCD prevalence and health inequalities of actionable changes in urban form; and (iv) develop an interactive toolkit that will support planners and policymakers responsible for delivering future urban form.

### Phase 1 – Building the evidence base on the relationship between urban design and NCDs

City-level urban design data derived from small-scale urban imagery, broadly consistent with Thompson et al. [36], will be selected for all UK and Australian cities with populations greater than 100,000. Areas selected for analysis will be based on agreed urban municipal boundaries for each city in Australia and the UK. This will capture approximately 90% of the UK and Australian populations.

Local-level image representations of each city will be gathered from a combination of maps, street-level, satellite and or aerial imagery where available. These images will be analysed for the presence of urban infrastructure and features informed by existing evidence and theory such as road networks and road types (e.g., local/ arterial/highway), intersections, rail transit networks, cycling networks, blue space, vehicles, people, and designated parks, tree canopies, tree types, and greenspace. Ideally, aerial and satellite imagery will have spatial resolution of < 25cm > 5cm. For context, at 20cm resolution we can visualise cars, and at 12.5cm resolution we can visualise individual people in the images. The utility of each perspective will be judged in the analytical phase against its association with population health outcomes, as well as its ease of integration into a computationally efficient workflow.

Figure 1 shows typical images proposed for our analysis, featuring map abstractions, satellite images, street view and sky-view. A grid-based dataset encompassing the entire sampling area for each city will be generated.

**Figure 1.**
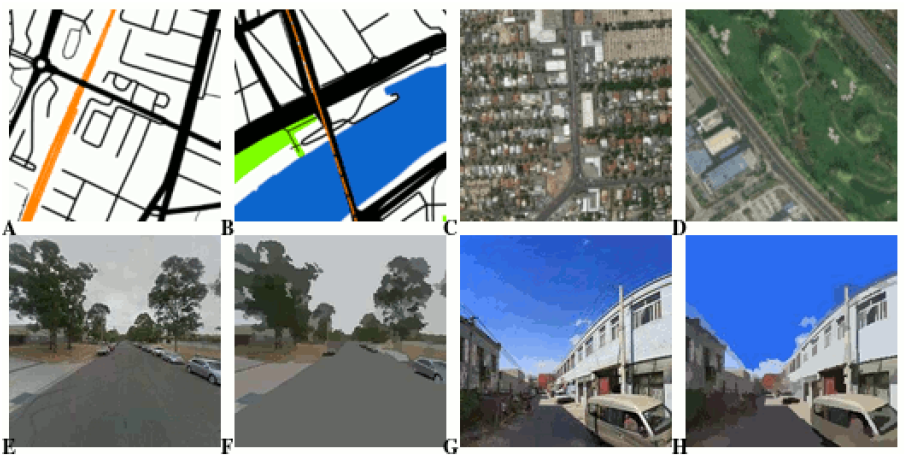
Four sample images featuring abstract maps (A,B), satellite view (C,D), street - view (E,F), and sky-view (G,H).

With the map image database for Australian and UK cities established, computer vision algorithms will be applied to the imagery dataset to identify given urban infrastructure features present in each image scaled to approximately 30m × 30m. These will include, but will not be limited to, footpaths, cycle paths, roads and road types, intersections, parks, sporting facilities, industrial complexes, shopping precincts, blue and green space, and other features identified through the literature.

Feature counts in these and surrounding images will then be undertaken using a raw or gravity model using the base image as the centroid and calculated for each image tile in each dataset with the exception of edge cases where the radius of the captured area around the centroid is curtailed.

In addition to features counted within each image and surrounding area, neighbourhoods will be classified based on the combination of features they contain and categorised into a total of ≥ 3 ≤ 10 groups using factor analysis or k-means clustering techniques.

A second method will also be employed whereby image data from all sampled locations across cities will be represented on a 2-dimensional self-organising map based on an adaptation of the city ‘fingerprint’ method of Nice and colleagues [37]. This multidimensional process will incorporate all available information collected through the feature identification methods, above and project each image or neighbourhood onto a standardised representation of the entire range of urban form observed in all cities in this research program from across the UK, Australia, or both. Following health data collection (see below) each position on the grid will also be matched to its performance across health risk and outcome measures while also controlling for socio-demographic differences between locations. The resulting ‘meta-map’ will provide a fine-grained representation of health risks and outcomes associated with unique combinations of observed urban design features, which can be matched back to any current or planned real-world location (Figure 2). The meta-map will: (i) form the definitive set of associations by which urban design can facilitate reductions across NCDs; and (ii) enable comparisons of health impact for populations between locations and given desired or planned changes in the characteristics of individual locations.

**Figure 2.**
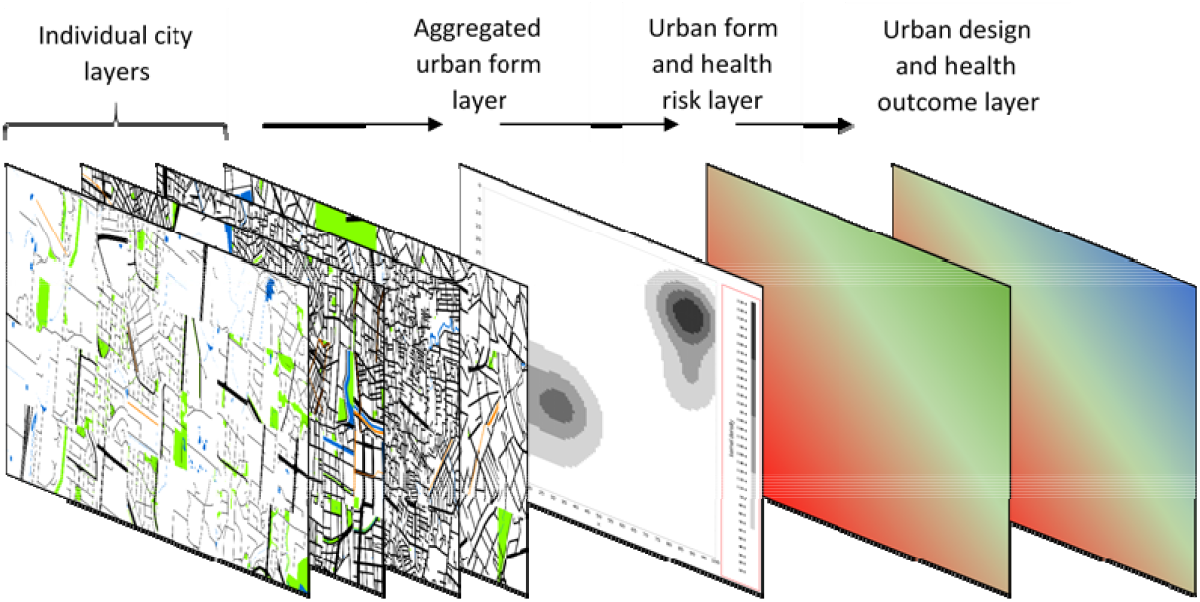
Representation of the city fingerprint and health impact process taking data from individual cities, combining all urban form into an aggregated ‘fingerprint’ map, and associating it with health risk and outcome data.

#### City types, health risk identification, and health outcomes

To investigate the association between city and within-city neighbourhood types and NCD risks and outcomes, available data from a range of sources aggregated to individual area levels will be utilised (e.g., beginning at the smallest available aggregated data-collection level such as Lower Layer Super Output Area (UK) or Statistical Area Level 2 (Australia) to calculate the risk for each NCD category for each city type or area, using the lowest risk type as the benchmark and controlling for city and national level confounders (e.g., economic indicators such as GDP or income, etc.). For health data, data from similar government sources in respective territories and jurisdictions will be sought (see Table 1 for examples).

**Table 1.**
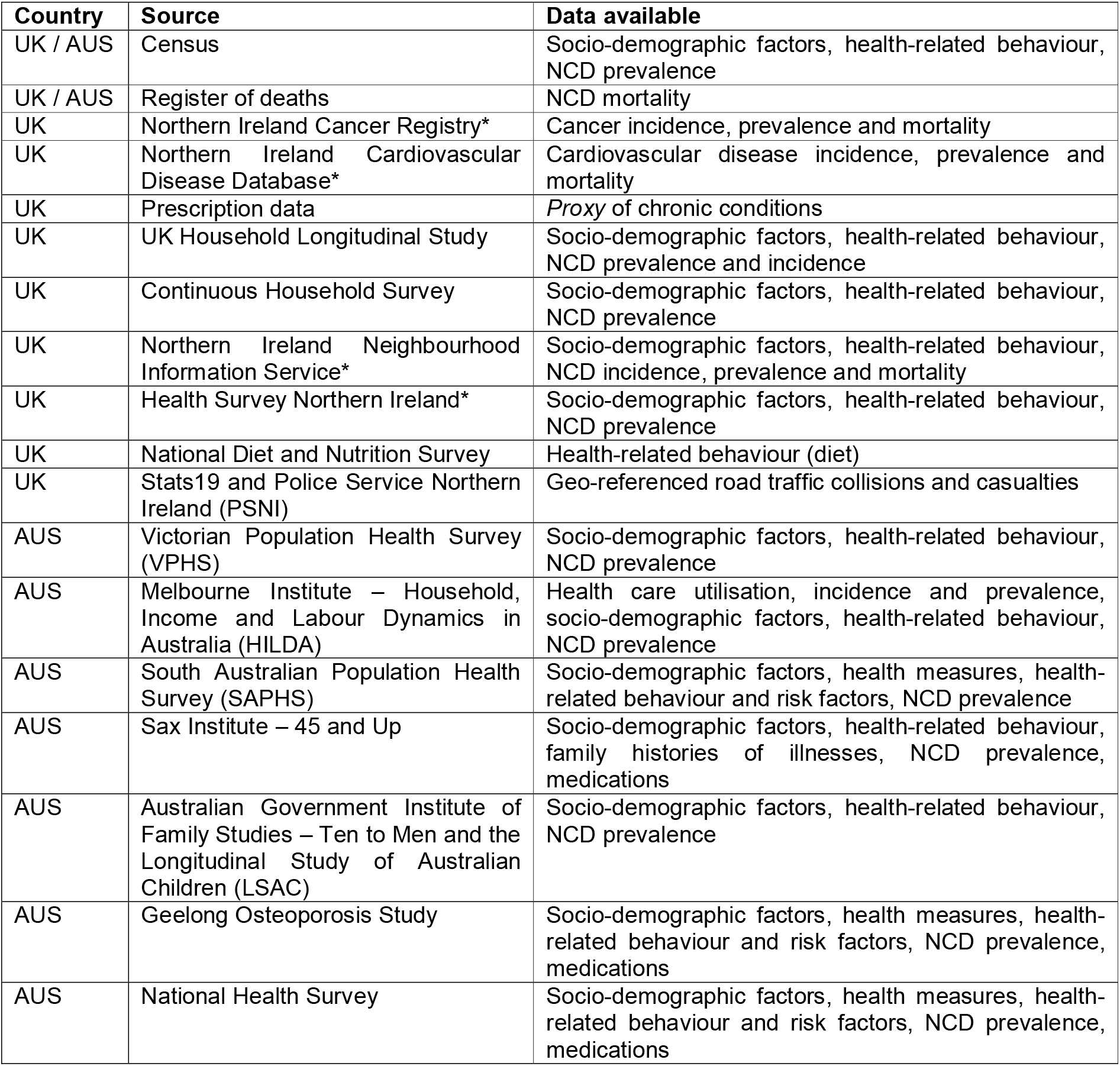
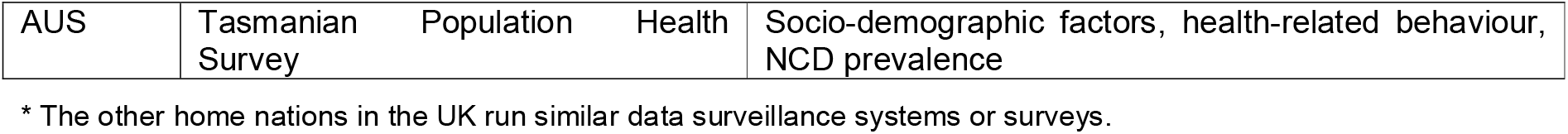
Example sources of geo-referenced health data available to the research team for analysis and comparison.

Road injury data will also be included in our analysis for two reasons. Firstly, road injury continues to be a significant global health burden, contributing to over 1.35 million deaths and 50 million injuries [46], ranking it as the 8th leading cause of death for people of all ages and the leading cause for children and young people aged 5-29 years of age. Secondly, our previous work modelling the nexus between urban design and health [16] indicates that urban design changes can reduce risk of selected NCDs but increase risk of road injury, compromising total health benefit [36]. This highlights complex interactions between urban design, NCDs, and road injury. For additional road transport and injury data, we will therefore access available geo-located road traffic collision and casualty data from state-based road transport authorities in Australia (e.g., Victorian Department of Transport) and in the UK from the Police Service Northern Ireland and Stats19 [47].

### Phase 2 - Investigating possible causal pathways

We will initially use health and GIS data from the UK Biobank [48], the 45 and Up study [49], and similar population health surveys in Australia, to prospectively investigate the possible causal pathways between urban design, NCD risk factors, and NCD incidence, and effect modifications associated with socio-demographic factors at the individual level. UK Biobank is a cohort of ∼500,000 adults aged 40-69 years from the general population, recruited between 2006 and 2010. It collects a wide range of socio-demographic (e.g., age, sex, ethnicity, education, and deprivation index) and health data (e.g., diet, physical activity, smoking, alcohol consumption, and NCD incidence) and provides objectively assessed built environment metrics from the immediate residential neighbourhood of UK Biobank participants, including building typology, destination accessibility, greenness, land use density, and street network accessibility [48]. The 45 and Up study data includes information from ∼250,000 participants from New South Wales, Australia, and will provide information on demographic and health-related factors such as body mass index, blood pressure, diet, levels of physical activity and physical energy expenditure for participants aged 45 years and above. GIS data, geocoded from home addresses, will also be provided by the 45 and Up data custodians.

Most statistical methods can only confirm associations between variables rather than causation. However, our research aims to explore the causal pathways through which urban design can affect NCD risk factors and NCD incidence and prevalence. Therefore, we will employ causal inference methods in this Phase.

Bayesian networks will be used to investigate multiple potential causal paths through which urban design can affect NCD risk factors and NCD incidence and prevalence. Bayesian networks are probabilistic directed acyclic graphs (DAGs) that can be used to obtain the statistical dependency structure between specific NCD risk factors and NCD incidence given different aspects of the built environment. DAGs first recover an undirected graph (i.e., a skeleton of the graph) to capture relationships between variables, while the direction of the identified relationships is determined in a subsequent step. DAGs are an established approach to investigate statistical dependency structures and have been successfully applied in a wide variety of domains, such as inferring gene interactions [50], assessing population health risks [51] and investigating tropical cyclone formation [52].

Also, we will compute changes in the conditional probabilities according to socio-demographic factors, offering a full and nuanced picture of whether and how social disparities moderate the relationship between built environment and NCD prevention. Each socio-demographic factor, built environment aspect, NCD risk factor and NCDs will be individually represented as a node in the DAGs. The DAGs structure will take into account prior evidence and knowledge (e.g., disallowing an NCD risk factor to be a parent node of sex or age), supplemented by applying learning algorithms to the observed data. Local conditional distributions will be derived from the data, and parameters will be obtained by performing Bayesian parameter estimation. Bootstrap resampling will be applied to reduce the impact of locally optimal networks on learning and inference. For purposes of validation, we will also utilise k-fold cross-validation [53].

### Phase 3 – Estimating health impacts of actionable changes in urban design

Building on Phases 1 and 2, this Phase will provide estimates of health impacts achievable via actionable changes in urban design. First, we will analyse the potential effects of city-wide changes in the built environment, benchmarking against healthier nearest-neighbour locations within the network graph generated in Phase 1. Second, we will investigate the range of urban morphology within each one of the sampled cities to provide a fine-grained representation of NCD risks and outcomes associated with unique urban design features, which can be used to suggest healthier solutions for existing areas and locations. This entire process will also provide a means to estimate potential reductions in within-city health inequalities driven by actionable changes in the urban morphology.

At the end of Phases 1 and 2, we will have metrics that allow us to estimate the changes in NCDs and other health outcomes (e.g., years of life lost due to premature death (YLL) and disability adjusted life-years (DALYs) averted) for a population based on changes to existing urban form or benefits associated with planned construction of new areas. As each city and geographic area will be identifiable, it will also highlight specific nearest-neighbour locations that can be used as feasible real-world examples to plan pathways for change from current to desired end-states (e.g., a desire to change from a high pollution, car-oriented urban network to the most feasibly achievable low pollution, pedestrian-friendly network). Therefore, for each location we can calculate changes in NCDs and health outcomes by comparing against: (i) healthier nearest-neighbour locations (assuming the location with the healthiest built environment as the benchmark); and (ii) the location with the healthiest built environment in the same country. The first approach will allow us to estimate the potential for NCD prevention in a realistic, short-term case scenario. The second approach allows estimation of the NCD prevention benefits associated with more ambitious but still achievable changes.

Complementarily, as in our previous work, generative adversarial networks will be used to identify and visualise optimal designs to transition the urban form from ‘unhealthy to healthy’ [38] and ‘‘unsafe to safe’ cycling [39] with resultant benefits in NCD prevention.

#### Supporting evidence-informed decisions for healthy urban design

##### Phase 4 - Developing a toolkit for action

We will develop a web-based tool, called City Vision, in which outputs from Phases 1-3 are presented and manipulated in an interactive environment, encouraging exploration of findings, new insights, and evidence-informed decisions. The toolkit will be co-designed with intended users (e.g., decision-makers and public servants in urban planning and health departments, members of the public, NGOs, researchers) to ensure that features required to match their tasks and goals are implemented, and that the tool can support their decisions and actions for NCD prevention. A usability study will be conducted alongside the tool development to identify the tasks and goals of intended users and the features required in the tool to match them and examine tool utility. It is anticipated that a group of 10-12 participants from the UK and Australia (invited from those living and working in the cities in Phase 1-3s), representing a variety of potential users, will offer critical insights and feedback on the structure, usability, and interface of the platform to meet the requirements of their decision-making cycle. Participants will be involved in three main activities: (i) in early stages of tool development, a one-day participatory workshop will discuss which features are required to meet their tasks and goals; (ii) during tool development, participants will test intermediate versions of the tool and provide feedback to improve features and overall usability; (iii) after completion of tool development, a second one-day meeting for participants to engage with the tool, discuss data gaps and priorities, determine what aspects of the process they found useful (or not) and why, discuss next steps to improve the tool and how it can be scaled up and applied to other settings. The research team will conduct and record the workshops, documenting decisions and outputs.

## DISCUSSION

We are living in a new urban age, an age awash with anthropogenic processes that influence health risks, but also, with new ways to understand them and potentially intervene. It is critical that we improve our understanding of the strengths and weaknesses of existing city designs to ensure they are safe, clean, healthy, and sustainable. Cities are dynamic – their infrastructure and people change, grow, adapt and develop habits and cultures, healthy or otherwise. Technologies advance and decline or are displaced. These technologies (e.g., the motor vehicle) create (in)efficiencies, solutions, bottlenecks, demands, and instigate new health and social challenges. There is no ideal healthy city that is impervious to change nor invulnerable to future, unforeseen, sociotechnical challenges.

Recent methodological advances in computer vision, AI, causal inference and complexity science as described in this research program offer opportunities to address long-standing research questions [42,45,54] about the relationship between urban design and health – and perhaps more importantly – what opportunities for positive change exist. We envisage facing some practical and technical challenges during the integration of these novel methods. For example, results might be influenced by unobserved or related factors such as historic patterns of social or urban development, or other conditions not captured by our observations but present within the health and imagery datasets. Secondly, there may be issues with the supply or harmonisation of datasets provided by different sources across countries – particularly in Australia which will require a consolidation and integration of a patchwork of health data.

The expected study benefits include: (i) a deeper understanding of how urban design and effective urban planning can prevent NCDs; (ii) how future programs and policies may better harness the power of the built environment to generate meaningful changes in NCD risk factors and consequent NCD incidence; and, (iii) identification of practical pathways for change using a nearest-neighbour strategy. Long term, benefits are expected to include the development of programs, policies and direct design and planning interventions that utilise the urban environment for NCD prevention and reduced risk, leading to better quality of life alongside longer life expectancy. Urban design and planning practitioners will benefit from a deeper understanding of how urban design and planning can reduce NCD risk factors and prevent NCDs in turn.

In summary, the future design of cities will be crucial to reducing the incidence, prevalence, and costs associated with NCDs. This study shows a new, replicable, scalable, and highly efficient method for understanding the characteristics of urban design with respect to NCDs and NCD risk factors. The research program highlights the opportunity for reducing the global burden of NCDs by embracing urban designs that emphasise characteristics captured within healthier city types.

## Data Availability

All data produced in the present study are available upon reasonable request to the authors

https://www.ukbiobank.ac.uk/

## ETHICS AND DISSEMINATION

The study was approved by the University of Melbourne, Human Research Ethics Committee (2021-20546-13629-3, approved on the 13/01/2021) and the Medicine, Health and Life Sciences Faculty Research Ethics Committee, Queen’s University Belfast (MHLS 20_85; 02/02/2021).

UK Biobank received ethical approval from the North West Multi-centre Research Ethics Committee (REC reference: 11/NW/03820). All participants gave written informed consent before enrolment in the study, which was conducted in accord with the principles of the Declaration of Helsinki.

